# Health insurance coverage, affordability barriers, and treatment gaps among Kenyan adults with diagnosed hypertension or diabetes before the Social Health Authority transition: a sex-stratified analysis of the 2022 Kenya Demographic and Health Survey

**DOI:** 10.64898/2026.05.14.26353157

**Authors:** Nichodemus Werre Amollo, Herieth F. Hyera, Ogol Japheth Ouma

**Author notes:** Correspondence: Nichodemus Werre Amollo.

## Abstract

**Background:** Kenya replaced the National Health Insurance Fund with the Social Health Authority in October 2024, making the 2022 Kenya Demographic and Health Survey the last nationally representative pre-transition baseline. Evidence on insurance coverage and treatment gaps among adults already diagnosed with hypertension or diabetes remains limited, including how these patterns differ by sex. We aimed to estimate the level, distribution, and correlates of insurance coverage and treatment gaps among diagnosed adults at the close of the NHIF era.

**Methods:** We conducted a survey-weighted cross-sectional secondary analysis of the 2022 Kenya Demographic and Health Survey. Insurance status, prior diagnosis, and current medication use were reported by respondents. Analyses were sex-stratified and survey-weighted, with adjusted prevalence ratios estimated to assess associations between insurance coverage and treatment gaps. Wealth-related inequality was examined using concentration indices.

**Results:** The analytic sample included 1,932 diagnosed adults (1,384 women and 548 men). Any insurance coverage was 47.7%, largely driven by National Health Insurance Fund enrolment (43.4%). Overall, 63.8% of diagnosed adults were classified as having a treatment gap (not currently taking medication for at least one diagnosed condition), including 67.1% of women and 59.8% of men, with treatment gaps exceeding 60% across all wealth quintiles. Insurance coverage was strongly pro-rich, whereas treatment gaps were distributed across the wealth gradient. After adjustment, insurance was not strongly associated with lower treatment-gap prevalence among women or men, and formal interaction testing did not support effect modification by sex. Among women, lack of money for treatment was reported as a major barrier far more frequently among the uninsured than the insured.

**Conclusions:** Before the Social Health Authority transition, Kenya faced unequal insurance coverage, persistent affordability barriers, and substantial treatment gaps among respondents diagnosed with hypertension or diabetes. These findings provide a national pre-transition benchmark for health financing reform. They suggest that expanding enrolment is necessary but unlikely to close chronic-care treatment gaps unless public insurance arrangements also improve benefit depth, outpatient medicine access, and frontline readiness for continuous NCD care.

## Background

Hypertension and diabetes are now leading drivers of avoidable morbidity, premature mortality, and household financial stress in Kenya. The 2015 Kenya STEPwise national survey estimated an adult hypertension prevalence of 24.5% and a diabetes prevalence of 2.4%, with awareness, treatment, and control levels well below targets for non-communicable diseases (NCDs) [1]. The Global Burden of Disease 2021 analysis confirms that diabetes continues to expand rapidly across sub-Saharan Africa [2]. For the health system, however, the more immediate policy question concerns adults who already know they have these conditions: whether they are financially protected and able to remain on treatment. That question became more urgent after Kenya replaced the National Health Insurance Fund (NHIF) with the Social Health Authority (SHA) on 1 October 2024 [3]. To date, nationally representative evidence on insurance coverage and treatment continuity among adults already diagnosed with hypertension or diabetes at the close of the NHIF era has remained limited, leaving the SHA reform without a credible pre-transition baseline.

In the general adult population, Kenya’s health insurance literature has consistently shown that coverage has remained low, socially patterned, and concentrated among better-off households. Using the 2008-09 Kenya Demographic and Health Survey (KDHS), Kimani et al. found strong socioeconomic gradients in women’s insurance ownership, particularly by wealth, education, and employment status [4]. Kazungu and Barasa later showed that insurance coverage in Kenya remained low and highly unequal even as coverage expanded between DHS waves, with a markedly pro-rich distribution [5]. At the wider sub-Saharan African level, Barasa et al. showed that insurance coverage is generally low and strongly wealth-skewed, especially where voluntary contributory mechanisms dominate [6]. Those studies are important, but they leave unresolved a closely related policy question: what was the insurance and treatment situation among adults already living with diagnosed hypertension or diabetes at the end of the NHIF era?

The Kenya NCD literature also leaves an important gap. Oyando et al. showed that NHIF provided some financial protection to households with hypertension and diabetes in a two-county study, but that evidence was not nationally representative and focused on household financial burden rather than national treatment coverage among diagnosed adults [7]. Recent mixed-methods work from rural primary health facilities in Kisumu documented medicine stock-outs, absent NCD budget lines, and structural financing barriers to effective hypertension and diabetes care [8]. Large treatment gaps in diagnosed populations can reflect incomplete access, discontinuation after treatment initiation, or imperfect adherence, so nationally representative evidence is needed on where diagnosed adults stood at the end of the NHIF era.

The 2022 KDHS offers a rare opportunity to examine that pattern nationally. It is the last nationally representative household survey conducted before the SHA transition, and it includes adult interview data on self-reported hypertension and diabetes diagnoses, current medication use, and household-member insurance coverage [9].

This study therefore aimed to estimate, using KDHS 2022, the level and distribution of insurance coverage and treatment gaps among adults who reported a prior clinician diagnosis of hypertension or diabetes in Kenya. Specifically, we examined three questions: (1) what proportion of diagnosed adults had any insurance and what proportion had NHIF coverage; (2) what proportion remained in a treatment gap; and (3) whether insurance coverage was associated with treatment gaps after accounting for sex, age, diagnosis profile, and socioeconomic position. We additionally quantified wealth-related inequality in insurance coverage and treatment gaps and examined women-specific care barriers to clarify whether financial protection and access barriers moved together.

## Methods

### Study design and data source

This study is reported in accordance with the Strengthening the Reporting of Observational Studies in Epidemiology (STROBE) statement; the completed checklist is provided as Additional file 2.

We conducted a cross-sectional secondary analysis of the 2022 Kenya Demographic and Health Survey (KDHS 2022), a nationally representative household survey implemented by the Kenya National Bureau of Statistics with technical support from The DHS Program [9]. KDHS 2022 used a stratified two-stage cluster design. Women aged 15-49 years were eligible for interview in selected households if they were usual residents or had slept in the household the night before the survey. Men aged 15-54 years were interviewed in a subsample of households [9]. The 2022 survey used both short and full questionnaire versions; the full questionnaire contained the health insurance, health expenditure, food security, and disability modules relevant to this project [9].

The 2022 KDHS protocol received ethical clearance from the Kenya Medical Research Institute Scientific and Ethics Review Unit (KEMRI/SERU) and the ICF Institutional Review Board, and all respondents provided informed consent during fieldwork [9]. The present study is a secondary analysis of de-identified, publicly available microdata obtained from The DHS Program; no new participant contact occurred.

### Analytic files and merge strategy

The full questionnaire was assigned to a pre-specified half of sampled households in KDHS 2022 and included the chronic-disease items used in the women’s analysis (KDHS routinely fields a more detailed health and care-access questionnaire to women aged 15–49 years, so several outcomes of interest are observable only in this subsample) [9].

The analytic base for women’s outcomes was the women’s individual recode file and for men’s outcomes the men’s recode file. Because the standard individual-file health-insurance variables are empty in KDHS 2022, insurance information was drawn from the household member (person) recode file and linked to each respondent using the standard DHS merge identifiers (cluster, household, and line number) [10, 11]. The interview files remained the analytic bases throughout. The analysis scripts and variable handling are available in the study repository.

### Study population

The target population was adults with a respondent-reported prior clinician diagnosis of hypertension and/or diabetes. For women, we restricted the analysis to respondents aged 18–49 years in the long-questionnaire subsample because the women’s chronic-disease items are structurally observed only in this subsample. We note here, and revisit in the limitations, that this design feature means women aged 50 years and above are not represented and that estimates for women should therefore be interpreted as applicable to the reproductive-age population captured by the standard women’s questionnaire. For men, we included respondents aged 18-54 years. We then retained respondents who reported a diagnosis of hypertension and/or diabetes and had non-missing information on insurance coverage and medication status for the diagnosed condition set. The final analytic sample comprised 1384 women and 548 men, for a pooled sample of 1932 diagnosed adults. For clarity, the women’s restriction reflects the KDHS full-questionnaire design rather than an ad hoc post-survey subsample, but it also means that sex-specific estimates are descriptive rather than strictly age-equivalent comparisons.

### Outcome definition

The primary outcome was the treatment gap among diagnosed adults. We defined a respondent as having a treatment gap if they reported a prior clinician diagnosis of hypertension and/or diabetes but were not currently taking medication for at least one diagnosed condition at the time of interview. This “diagnosed-not-on-treatment” construct corresponds to the “treatment” step routinely used in hypertension- and diabetes-care cascade studies of nationally representative DHS- and STEPS-type surveys [10, 11]. For both sexes, hypertension and diabetes diagnoses were identified from the chronic-disease questions in the long-questionnaire module, and current medication use for each diagnosed condition was identified from the parallel medication-use questions. We also classified respondents into three diagnosis profiles: hypertension only, diabetes only, and both hypertension and diabetes. The primary outcome captures incomplete treatment, classifying a respondent as having a gap whenever at least one diagnosed condition is untreated; this is the clinically meaningful target for complete chronic-care coverage among adults with one or more diagnosed NCDs. As a sensitivity analysis, we additionally defined a stricter no-treatment outcome under which a respondent is classified as having a gap only when no medication is taken for any diagnosed condition; this stricter definition reduces the contribution of partially treated comorbid respondents and tests the robustness of the headline estimate. We therefore estimated this stricter no-treatment outcome alongside the primary outcome and compared the two.

### Insurance exposure and covariates

The primary exposure was any health insurance coverage, derived from the household roster insurance question. NHIF coverage was derived from the corresponding insurance-type item on the same roster. Because the insurance-type items are asked conditional on having insurance, NHIF was recoded as zero for respondents reporting no insurance and retained as missing only when insurance status itself was missing. We also generated descriptive indicators for private and community-based coverage, although NHIF was the dominant insurance type in this population. Insurance status came from the household roster rather than the interview files, so some misclassification remains possible if roster responses were inaccurate, although the roster and medication items both reflect current-status reporting in the same survey round.

Covariates were selected a priori from prior Kenya insurance and NCD literature [4–8]. These included age in years, sex, wealth quintile, educational attainment, place of residence, province, employment category, and diagnosis profile. Employment categories were collapsed to reduce sparse cells in regression models. For women, we additionally examined three access-barrier items from the standard DHS care-access module: obtaining permission to seek care, getting money for treatment, and distance to a health facility. Each was coded as “a big problem” versus “not a big problem or no problem.” The money-for-treatment item is reported by respondents in Kenyan shillings/local currency framing, but is captured as a perceived barrier rather than as a specific monetary amount, so no currency conversion to United States dollars (USD) is applicable; where monetary amounts appear elsewhere in this manuscript they are accompanied by USD equivalents to aid global comparability.

### Survey weighting and statistical analysis

Analyses respected the DHS complex sample design. Women’s models used the women’s interview weight (v005) with primary sampling units (v021) and strata (v022). Men’s models used the men’s interview weight (mv005) with design variables mv021 and mv022. All weights were divided by 1,000,000, consistent with DHS guidance and the Guide to DHS Statistics [12, 13]. Because men were interviewed in a household subsample, pooled descriptive estimates and pooled secondary regression models used a male-subsample correction derived within strata from the PR file, following DHS guidance on pooling male and female files when the men’s sample is subsampled [13, 14]. Primary inference remained sex-stratified.

Because the women’s chronic-disease module was fielded in the pre-specified full-questionnaire half-sample, women in eligible full-questionnaire households retained the standard DHS women’s weight after restriction [9, 14].

We first described the sample using weighted proportions and means with 95% confidence intervals. We then constructed sex-specific and pooled treatment-cascade tables summarising the prevalence of any insurance, NHIF coverage, and treatment gaps across diagnosis profiles. To quantify wealth-related inequality, we estimated concentration indices [15], a standard summary measure of socioeconomic-rank-related inequality in a health outcome, calculated as twice the area between the cumulative-population concentration curve and the line of equality. Positive values indicate that the outcome is more concentrated among wealthier individuals; negative values indicate concentration among poorer individuals. We obtained these for insurance coverage, NHIF coverage, and treatment gaps with bootstrap uncertainty intervals [15, 16].

We report the standard concentration index in the main text for comparability with prior Kenya inequality studies and estimated Erreygers-corrected indices as sensitivity analyses because insurance coverage and treatment gaps were binary outcomes [15, 16]. All concentration-index uncertainty intervals were based on 1,000 bootstrap replications.

For regression analyses, we estimated survey-weighted generalized linear models with a quasi-Poisson family and log link to obtain adjusted prevalence ratios (APR) for the cross-sectional association between insurance coverage and treatment gaps [17]. Prevalence ratios were preferred over odds ratios because treatment gaps were common (well above 10%) and odds ratios can exaggerate the apparent magnitude of association for common outcomes. All primary models were fitted separately for women and men because the women’s and men’s questionnaires are not interchangeable in KDHS and women aged 50–54 years are not observed. A pooled model with a sex term was retained as a secondary analysis after correcting men’s weights for subsampling. Survey-weighted logistic models were fitted as odds-ratio sensitivity analyses and are reported in Additional file 1: Supplementary Table S1. We repeated the prevalence and regression analyses using the stricter no-treatment definition; those results are reported in Additional file 3: Supplementary Table S2.

Secondary pooled models also tested insurance-by-sex and sex-by-wealth interaction terms using design-based Wald tests. Because current insurance and current medication use were measured contemporaneously, the models were not intended to establish temporal ordering or causality.

All analyses were performed in R 4.5.1 using the survey package (version 4.4.2) for complex sample estimation [17]. Final concentration indices were estimated with a custom bootstrap-based weighted-rank routine implemented in R [15, 16]. Full analysis scripts are available at https://github.com/gondamol/Kenya_DHS_Studies/tree/main/ST03_NCD_Insurance_Service_Use.

## Results

### Sample characteristics and insurance profile

The final analytic sample included 1932 adults with a respondent-reported prior clinician diagnosis of hypertension and/or diabetes, of whom 1384 were women and 548 were men. Mean age was 35.5 years among women, 40.5 years among men, and 37.8 years in the pooled sample (Table 1). Weighted any-insurance coverage was 42.7% (95% CI 39.2% to 46.2%) among women, 53.7% (95% CI 48.4% to 59.0%) among men, and 47.7% (95% CI 44.4% to 50.9%) overall. NHIF remained the dominant form of coverage: pooled NHIF coverage among all diagnosed adults was 43.4% (95% CI 40.1% to 46.7%), while NHIF covered 91.1% (95% CI 88.0% to 94.1%) of insured diagnosed adults.

**Table 1.** Sample characteristics among diagnosed adults with hypertension and/or diabetes, Kenya DHS 2022.

| Characteristic | Women |  | Men |  | Pooled |  |
| --- | --- | --- | --- | --- | --- | --- |
|  | n | Estimate (95% CI) | n | Estimate (95% CI) | n | Estimate (95% CI) |
| Sample size, n | 1,384 |  | 548 |  | 1,932 |  |
| Mean age, years (95% CI) | 1,384 | 35.5 (35.0, 36.1) | 548 | 40.5 (39.3, 41.7) | 1,932 | 37.8 (37.1, 38.4) |
| Diagnosis profile |  |  |  |  |  |  |
| HTN only | 1,258 | 90.9 (88.4, 93.3) | 412 | 74.9 (69.6, 80.2) | 1,670 | 83.7 (81.0, 86.5) |
| DM only | 85 | 5.8 (4.1, 7.5) | 87 | 13.2 (9.6, 16.8) | 172 | 9.2 (7.3, 11.0) |
| Both HTN and DM | 41 | 3.3 (1.5, 5.2) | 49 | 11.9 (7.2, 16.5) | 90 | 7.1 (4.8, 9.4) |
| Wealth quintile |  |  |  |  |  |  |
| Poorest | 163 | 8.9 (7.1, 10.7) | 68 | 9.9 (7.2, 12.6) | 231 | 9.5 (7.7, 11.2) |
| Poorer | 205 | 13.1 (11.0, 15.1) | 81 | 13.1 (9.9, 16.3) | 286 | 13.2 (11.3, 15.1) |
| Middle | 302 | 19.2 (16.7, 21.6) | 113 | 18.6 (15.1, 22.0) | 415 | 18.9 (16.7, 21.1) |
| Richer | 355 | 24.1 (20.8, 27.4) | 141 | 24.1 (19.2, 29.1) | 496 | 24.0 (21.0, 27.1) |
| Richest | 359 | 34.7 (30.0, 39.4) | 145 | 34.3 (29.1, 39.5) | 504 | 34.4 (30.6, 38.2) |
| Educational attainment |  |  |  |  |  |  |
| None or primary | 692 | 44.7 (40.6, 48.8) | 209 | 35.6 (30.3, 40.9) | 901 | 40.6 (37.2, 44.0) |
| Secondary+ | 692 | 55.3 (51.2, 59.4) | 339 | 64.4 (59.1, 69.7) | 1,031 | 59.4 (56.0, 62.8) |
| Residence |  |  |  |  |  |  |
| Urban | 601 | 46.6 (43.2, 50.1) | 224 | 42.8 (39.9, 45.7) | 825 | 44.9 (42.0, 47.9) |
| Rural | 783 | 53.4 (49.9, 56.8) | 324 | 57.2 (54.3, 60.1) | 1,107 | 55.1 (52.1, 58.0) |
| Employment |  |  |  |  |  |  |
| Agriculture | 240 | 16.7 (14.2, 19.2) | 139 | 23.7 (19.8, 27.5) | 379 | 19.9 (17.5, 22.3) |
| Professional/clerical | 302 | 22.0 (19.1, 25.0) | 130 | 22.5 (17.4, 27.5) | 432 | 22.2 (19.4, 25.1) |
| Sales/services | 300 | 24.9 (21.1, 28.6) | 84 | 16.7 (11.5, 22.0) | 384 | 21.3 (18.2, 24.3) |
| Manual | 164 | 11.8 (9.4, 14.1) | 146 | 26.5 (21.2, 31.8) | 310 | 18.2 (15.5, 21.0) |
| Not working | 378 | 24.6 (21.1, 28.2) | 49 | 10.6 (5.8, 15.5) | 427 | 18.3 (15.5, 21.1) |
| Any insurance | 1,384 | 42.7 (39.2, 46.2) | 548 | 53.7 (48.4, 59.0) | 1,932 | 47.7 (44.4, 50.9) |
| NHIF coverage | 1,384 | 38.4 (34.8, 42.1) | 548 | 49.5 (44.4, 54.7) | 1,932 | 43.4 (40.1, 46.7) |
| Treatment gap | 1,384 | 67.1 (63.5, 70.6) | 548 | 59.8 (53.8, 65.8) | 1,932 | 63.8 (60.4, 67.3) |

Source: Kenya DHS 2022. Survey-weighted estimates.

**Table 2.** Insurance coverage and treatment gaps by diagnosis profile and sex, Kenya DHS 2022.

| Sex | Diagnosis profile | n | Any insurance<br>% (95% CI) | NHIF coverage<br>% (95% CI) | Treatment gap<br>% (95% CI) |
| --- | --- | --- | --- | --- | --- |
| Women | HTN only | 1,258 | 42.4 (38.8, 46.1) | 37.9 (34.3, 41.5) | 68.9 (65.2, 72.7) |
|  | DM only | 85 | 42.2 (25.7, 58.8) | 40.9 (24.3, 57.4) | 46.5 (31.9, 61.0) |
|  | Both HTN and DM | 41 | 50.1 (14.5, 85.7) | 50.1 (14.5, 85.7) | 51.9 (15.8, 87.9) |
|  | Any diagnosed NCD | 1,384 | 42.7 (39.2, 46.2) | 38.4 (34.8, 42.1) | 67.1 (63.5, 70.6) |
| Men | HTN only | 412 | 51.8 (45.9, 57.7) | 48.0 (42.3, 53.7) | 69.1 (62.3, 75.9) |
|  | DM only | 87 | 44.9 (32.4, 57.3) | 36.2 (19.8, 52.5) | 31.2 (15.3, 47.1) |
|  | Both HTN and DM | 49 | 75.8 (64.6, 86.9) | 74.4 (63.1, 85.7) | 32.9 (18.5, 47.2) |
|  | Any diagnosed NCD | 548 | 53.7 (48.4, 59.0) | 49.5 (44.4, 54.7) | 59.8 (53.8, 65.8) |
| Pooled | HTN only | 1,670 | 46.3 (42.8, 49.7) | 41.9 (38.5, 45.3) | 69.1 (65.5, 72.6) |
|  | DM only | 172 | 43.6 (33.6, 53.6) | 37.6 (26.8, 48.4) | 36.3 (25.3, 47.2) |
|  | Both HTN and DM | 90 | 69.2 (55.3, 83.1) | 68.1 (54.2, 82.1) | 37.9 (23.2, 52.7) |
|  | Any diagnosed NCD | 1,932 | 47.7 (44.4, 50.9) | 43.4 (40.1, 46.7) | 63.8 (60.4, 67.3) |

Source: Kenya DHS 2022. Survey-weighted estimates.
Estimates for 'Both HTN and DM' should be interpreted with caution owing to small unweighted sample sizes (women n=41, men n=49). Wide confidence intervals reflect sampling imprecision.

**Table 3.** Adjusted prevalence ratios for the treatment gap among diagnosed adults, Kenya DHS 2022.

| Covariate | Women<br>APR (95% CI) | Men<br>APR (95% CI) | Pooled<br>APR (95% CI) |
| --- | --- | --- | --- |
| Insurance |  |  |  |
| Uninsured (ref=1.00) | 1.00 (ref) | 1.00 (ref) | 1.00 (ref) |
| Insured | 0.91 (0.81, 1.03) | 0.93 (0.75, 1.16) | 0.93 (0.83, 1.04) |
| Age (per year) | 0.99 (0.98, 0.99) | 0.99 (0.98, 1.00) | 0.99 (0.98, 0.99) |
| Diagnosis |  |  |  |
| Both HTN+DM (ref=1.00) | 1.00 (ref) | 1.00 (ref) | 1.00 (ref) |
| HTN only | 1.22 (0.68, 2.20) | 1.96 (1.23, 3.15) | 1.65 (1.12, 2.44) |
| DM only | 0.83 (0.43, 1.60) | 0.84 (0.43, 1.63) | 0.85 (0.52, 1.40) |
| Wealth |  |  |  |
| Richest (ref=1.00) | 1.00 (ref) | 1.00 (ref) | 1.00 (ref) |
| Richer | 0.92 (0.80, 1.07) | 0.95 (0.73, 1.24) | 0.94 (0.81, 1.09) |
| Middle | 0.94 (0.80, 1.10) | 0.85 (0.62, 1.17) | 0.90 (0.77, 1.06) |
| Poorer | 0.93 (0.78, 1.12) | 0.92 (0.65, 1.30) | 0.94 (0.79, 1.12) |
| Poorest | 0.86 (0.68, 1.08) | 0.95 (0.65, 1.38) | 0.92 (0.74, 1.13) |
| Education |  |  |  |
| None (ref=1.00) | 1.00 (ref) | 1.00 (ref) | 1.00 (ref) |
| Primary | 0.84 (0.67, 1.06) | 0.84 (0.59, 1.20) | 0.81 (0.65, 1.00) |
| Secondary+ | 0.88 (0.69, 1.12) | 0.80 (0.56, 1.15) | 0.82 (0.65, 1.02) |
| Residence |  |  |  |
| Rural (ref=1.00) | 1.00 (ref) | 1.00 (ref) | 1.00 (ref) |
| Urban | 0.98 (0.86, 1.11) | 0.96 (0.76, 1.22) | 0.98 (0.86, 1.10) |
| Employment |  |  |  |
| Agriculture (ref=1.00) | 1.00 (ref) | 1.00 (ref) | 1.00 (ref) |
| Professional/clerical | 0.92 (0.78, 1.08) | 0.90 (0.68, 1.18) | 0.92 (0.79, 1.07) |
| Sales/services | 0.81 (0.67, 0.98) | 0.72 (0.53, 1.00) | 0.78 (0.66, 0.91) |
| Manual | 1.02 (0.87, 1.20) | 0.82 (0.67, 1.01) | 0.92 (0.80, 1.05) |
| Not working | 0.88 (0.77, 1.01) | 0.69 (0.48, 1.00) | 0.83 (0.72, 0.95) |
| Province |  |  |  |
| Central (ref=1.00) | 1.00 (ref) | 1.00 (ref) | 1.00 (ref) |
| Coast | 0.84 (0.69, 1.01) | 0.70 (0.51, 0.95) | 0.78 (0.66, 0.93) |
| Eastern | 0.95 (0.82, 1.09) | 0.93 (0.70, 1.23) | 0.95 (0.83, 1.08) |
| Nairobi | 0.75 (0.57, 0.99) | 0.89 (0.55, 1.43) | 0.81 (0.63, 1.05) |
| NE | 0.89 (0.71, 1.13) | 0.61 (0.31, 1.21) | 0.73 (0.53, 0.99) |
| Nyanza | 0.79 (0.67, 0.93) | 1.06 (0.84, 1.34) | 0.92 (0.81, 1.06) |
| Rift Valley | 0.93 (0.82, 1.05) | 0.87 (0.67, 1.14) | 0.92 (0.81, 1.04) |
| Western | 0.88 (0.75, 1.04) | 0.72 (0.50, 1.02) | 0.82 (0.68, 0.98) |

Source: Kenya DHS 2022. Survey-weighted estimates. APR from survey-weighted quasi-Poisson GLM with log link. Reference categories are shown in parentheses. The pooled model includes a sex term with male-subsample correction. NE = North Eastern province.

### Treatment cascade by diagnosis profile

Treatment gaps were large in both sexes. Weighted treatment-gap prevalence was 67.1% (95% CI 63.5% to 70.6%) among women, 59.8% (95% CI 53.8% to 65.8%) among men, and 63.8% (95% CI 60.4% to 67.3%) overall.

Diagnosis-specific patterns showed substantial heterogeneity. Among women, the treatment gap was highest for hypertension only (68.9% (95% CI 65.2% to 72.7%)) and lower for diabetes only (46.5% (95% CI 31.9% to 61.0%)). Among men, the treatment gap was also highest for hypertension only (69.1% (95% CI 62.3% to 75.9%)), whereas men reporting both hypertension and diabetes had substantially higher insurance coverage (75.8% (95% CI 64.6% to 86.9%)) and lower treatment gaps (32.9% (95% CI 18.5% to 47.2%)). The pooled cascade showed the same pattern: treatment gaps were highest among respondents with hypertension only and lower among those with both conditions. Within the pooled comorbid subgroup, 25 respondents had a treatment gap in one condition and 24 had a gap in both conditions, while 41 reported current medication for both conditions. The diabetes-only and comorbid subgroups were small after sex stratification, so those subgroup-specific estimates should be interpreted cautiously.

### Adjusted associations with treatment gaps

In adjusted prevalence-ratio models, any insurance showed no strong association with treatment-gap prevalence in either sex. The APR for insured versus uninsured adults was 0.91 (95% CI 0.81 to 1.03) among women, 0.93 (95% CI 0.75 to 1.16) among men, and 0.93 (95% CI 0.83 to 1.04) in the pooled secondary model. Older age was associated with slightly lower treatment-gap prevalence in women, men, and pooled analyses. In the pooled model, adults with hypertension only had higher treatment-gap prevalence than adults with both hypertension and diabetes (APR 1.65, 95% CI 1.12 to 2.44). Treatment gaps were lower among adults working in sales and services than among those in agriculture, and several regional contrasts also persisted after adjustment, including lower pooled treatment gaps in Coast, North Eastern, and Western provinces relative to Central. Secondary interaction tests found no evidence that the association between insurance and treatment gaps differed by sex (p = 0.723) or that wealth gradients differed materially by sex (p = 0.675). Under the stricter sensitivity definition requiring no medication for any diagnosed condition, pooled no-treatment prevalence was 62.2% (95% CI 58.7% to 65.7%) and the adjusted pooled insurance association remained similar (APR 0.94 (95% CI 0.85 to 1.04); Additional file 3: Supplementary Table S2).

Odds-ratio sensitivity analyses are reported in Additional file 1: Supplementary Table S1. The stricter no-medication-for-any-diagnosed-condition sensitivity analysis is reported in Additional file 3: Supplementary Table S2. Directional conclusions were unchanged.

### Wealth-related inequality and women-specific access barriers

Insurance coverage among diagnosed adults was strongly pro-rich. The pooled standard concentration index was 0.2889 (95% CI 0.2341 to 0.2969) for any insurance and 0.2848 (95% CI 0.2232 to 0.2966) for NHIF coverage. By contrast, the standard concentration index for treatment gaps was −0.0347 (95% CI −0.0552 to 0.0066), suggesting that treatment gaps were widespread across the socioeconomic distribution rather than concentrated only among the poorest. Erreygers-corrected sensitivity analyses showed the same directional pattern (0.5507 (95% CI 0.4418 to 0.5687) for any insurance and −0.0887 (95% CI −0.1387 to 0.0174) for treatment gaps) (Table 4).

**Table 4.** Standard and Erreygers-corrected concentration indices for insurance coverage and treatment gaps among diagnosed adults, Kenya DHS 2022.

| Outcome | Subgroup | Standard CI<br>(95% CI) | Erreygers-corrected CI<br>(95% CI) |
| --- | --- | --- | --- |
| Any insurance | Pooled | 0.2889 (0.2341, 0.2969) | 0.5507 (0.4418, 0.5687) |
| NHIF coverage | Pooled | 0.2848 (0.2232, 0.2966) | 0.4944 (0.3784, 0.5163) |
| Treatment gap | Pooled | $-0.0347$ ($-0.0552$, $0.0066$) | $-0.0887$ ($-0.1387$, $0.0174$) |
| Any insurance | Women | 0.2636 (0.2188, 0.2991) | 0.4499 (0.3642, 0.5238) |
| Any insurance | Men | 0.2944 (0.2205, 0.3226) | 0.6326 (0.4754, 0.6812) |

Source: Kenya DHS 2022. Survey-weighted estimates.
Standard concentration indices are shown for comparability with prior Kenya inequality studies. Erreygers-corrected indices are included as sensitivity analyses for binary outcomes.

**Table 5.** Women-specific access barriers by insurance status among diagnosed adults, Kenya DHS 2022.

| Insurance type | Covered | Barrier type | n | Weighted prevalence % (95% CI) |
| --- | --- | --- | --- | --- |
| Any insurance | No | Money for treatment | 842 | 63.2% (95% CI 58.9% to 67.5%) |
| Any insurance | Yes | Money for treatment | 542 | 30.6% (95% CI 25.0% to 36.3%) |
| Any insurance | No | Distance to facility | 842 | 32.1% (95% CI 27.4% to 36.8%) |
| Any insurance | Yes | Distance to facility | 542 | 16.1% (95% CI 12.4% to 19.9%) |
| Any insurance | No | Permission to seek care | 842 | 6.4% (95% CI 4.0% to 8.8%) |
| Any insurance | Yes | Permission to seek care | 542 | 3.1% (95% CI 1.8% to 4.3%) |
| NHIF coverage | No | Money for treatment | 881 | 60.4% (95% CI 55.4% to 65.3%) |
| NHIF coverage | Yes | Money for treatment | 503 | 31.6% (95% CI 26.2% to 37.0%) |
| NHIF coverage | No | Distance to facility | 881 | 30.8% (95% CI 26.5% to 35.1%) |
| NHIF coverage | Yes | Distance to facility | 503 | 16.5% (95% CI 12.5% to 20.5%) |
| NHIF coverage | No | Permission to seek care | 881 | 6.1% (95% CI 3.9% to 8.3%) |
| NHIF coverage | Yes | Permission to seek care | 503 | 3.1% (95% CI 1.7% to 4.5%) |

Source: Kenya DHS 2022. Survey-weighted estimates.

Figure 1 illustrates these patterns across wealth quintiles. Insurance coverage rose steeply with wealth in both sexes, with men consistently more covered than women at every quintile and the gap between them widening in the upper distribution. By contrast, treatment-gap prevalence ran nearly flat — ranging from 69.0% (95% CI 62.5% to 75.6%) in the poorer quintile to 60.5% (95% CI 53.1% to 67.9%) in the richest — with confidence intervals for men and women overlapping at most quintiles.

**Figure 1.**
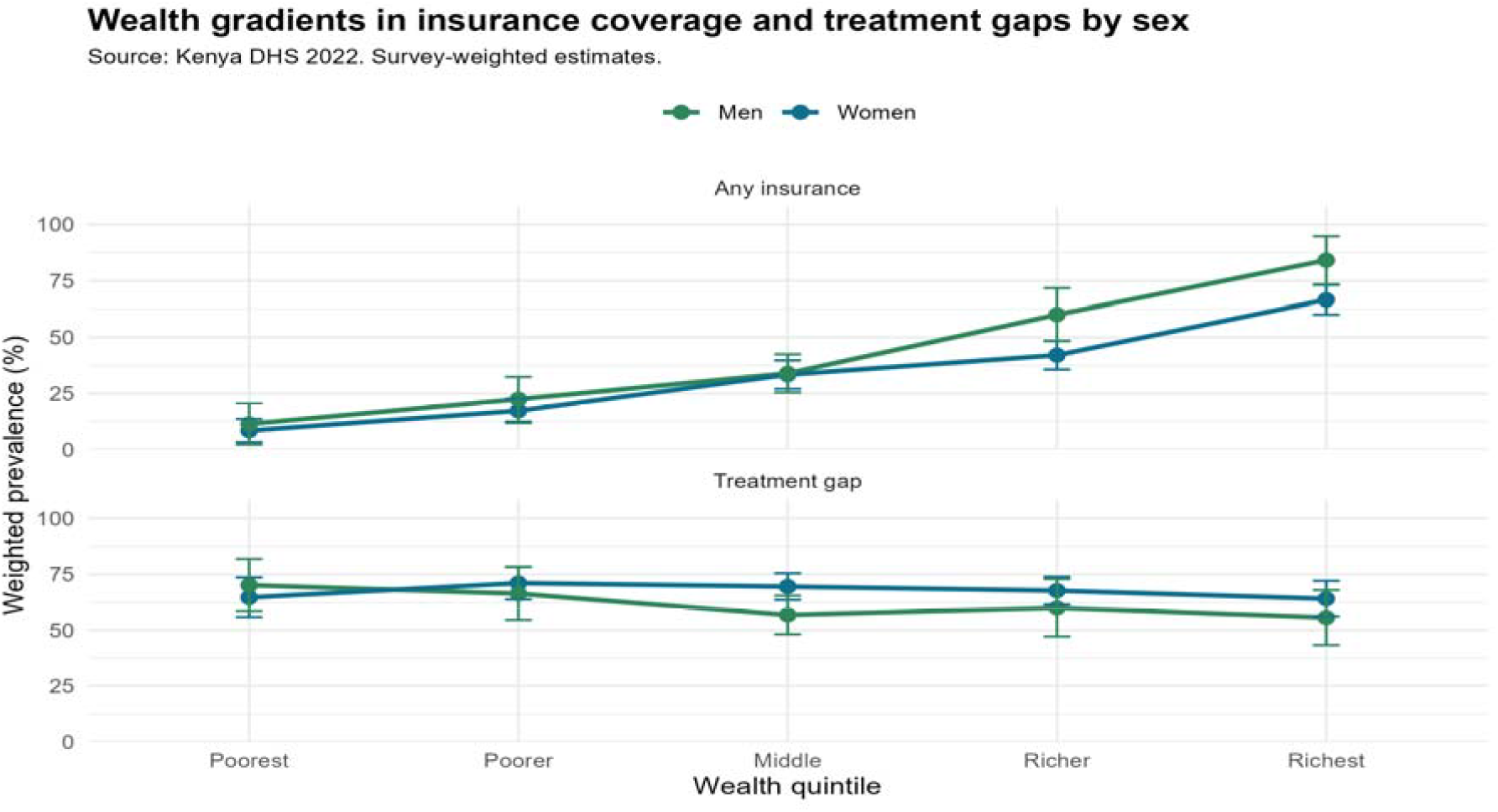
Wealth gradients in insurance coverage and treatment gaps by sex among respondents with diagnosed hypertension or diabetes, Kenya DHS 2022. Points and lines show survey-weighted proportions across wealth quintiles, stratified by sex. Error bars are 95% confidence intervals.

**Figure 1:**
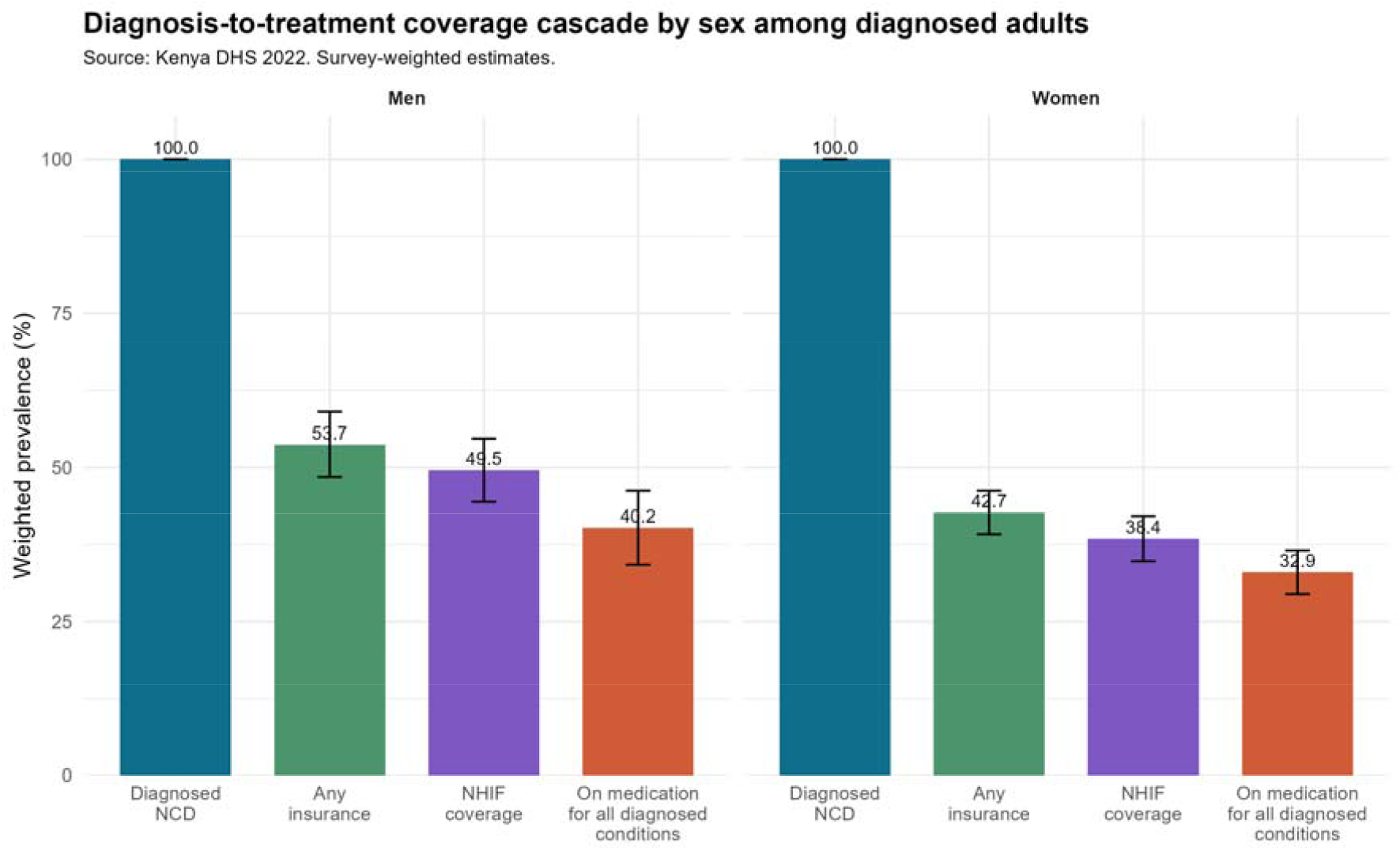
Diagnosis-to-treatment coverage cascade by sex among adults with self-reported diagnosed hypertension or diabetes, Kenya DHS 2022. Bars show the diagnosed population baseline (100%), any insurance coverage, NHIF coverage, and current medication use for

Among women with diagnosed hypertension and/or diabetes, financial (money for treatment) and geographic (distance to a health facility) access barriers were much more common among the uninsured than the insured. Reporting “a big problem” getting money for treatment was reported by 63.2% (95% CI 58.9% to 67.5%) of uninsured women versus 30.6% (95% CI 25.0% to 36.3%) of insured women. Distance to a health facility was a big problem for 32.1% (95% CI 27.4% to 36.8%) of uninsured women compared with 16.1% (95% CI 12.4% to 19.9%) of insured women. Permission to seek care was less common overall but remained more frequent among uninsured women (6.4% (95% CI 4.0% to 8.8%) versus 3.1% (95% CI 1.8% to 4.3%)). These barrier contrasts are descriptive and women-specific; they indicate perceived access constraints but are not equivalent to adjusted estimates of current medication use.

### Overall diagnosis-to-treatment coverage cascade

Figure 2 reframes the same problem as a diagnosis-to-treatment cascade. Among respondents who had already received a diagnosis, coverage fell from diagnosis to insurance and again from insurance to being on medication for all diagnosed conditions. The cumulative drop from diagnosis to current treatment was larger among women than men.

## Discussion

This study provides a national pre-SHA baseline for insurance coverage and treatment access among adults in Kenya who reported a prior clinician diagnosis of hypertension or diabetes. Three findings stand out. First, treatment gaps among diagnosed adults were substantial in both sexes. Second, insurance enrolment was incomplete, with fewer than half of diagnosed adults covered. Third, insurance status was not strongly associated with lower treatment-gap prevalence after adjustment. Together, these findings indicate that, by the close of the NHIF era, Kenya faced two interlinked problems in chronic care: incomplete enrolment in a fragmented insurance landscape, and incomplete effective coverage even where enrolment had been achieved.

The treatment gap is not a marginal finding. Nearly two in three adults who reported having already been told they had hypertension or diabetes were not taking medication for all reported diagnosed conditions. Despite growing evidence of widespread hypertension in sub-Saharan Africa, including a large burden of undiagnosed cases [18], awareness of a diagnosis does not reliably translate into sustained treatment uptake [19]. Comparable multi-country cascade analyses have documented similarly large losses between diagnosis and treatment for hypertension and diabetes across low- and middle-income countries [10, 11]. Because all respondents in our analytic sample reported a prior diagnosis, the observed gaps are unlikely to reflect screening failure alone. They more plausibly reflect some combination of interrupted access, discontinuation after treatment initiation, inconsistent adherence, or prescribing gaps. The stricter no-treatment sensitivity definition yielded a similar headline prevalence, which suggests that the headline finding is not driven solely by partial treatment among respondents with multiple diagnoses.

The weak adjusted association between insurance and treatment gaps is equally important. Insurance still mattered descriptively, particularly for women-specific affordability barriers, but current enrolment status alone was not accompanied by a strong difference in current medication use after adjustment. The concept of effective coverage, formalised by Shengelia et al. (2005), emphasises that service contact and financial coverage matter only when care of sufficient quality is actually received [20]. Our findings are consistent with this framework: adults may hold insurance cards yet still face medicine stock-outs, transport costs, user charges outside reimbursed services, or benefit-package arrangements that do not reliably translate into medicine access at the point of care. That interpretation is consistent with Kenya evidence on effective coverage [21], NHIF purchasing reforms [22, 23], global evidence on catastrophic household expenditure [24], and persistent catastrophic health expenditure among Kenyan households [25–27]. Recent synthesis work on hypertension and diabetes care cascades, together with the broader adherence literature, likewise suggests that treatment-stage losses reflect both health-system failures and long-term medication-taking behaviour rather than insurance status alone [28, 29]. At the same time, the data cannot distinguish whether untreated respondents lacked prescriptions, could not obtain medicines, discontinued treatment, or chose not to take it, so access failure and non-adherence should both remain plausible explanations. The contrast between the large women-specific barrier differences and the weak adjusted insurance association is therefore not internally inconsistent: the barrier items capture self-reported affordability and travel constraints among women only, whereas the regression models estimate the net cross-sectional association between insurance status and current medication use across both sexes after adjustment for diagnosis profile and socioeconomic position.

This interpretation also fits prior Kenya evidence. Oyando et al. reported that NHIF offered some financial protection for households affected by hypertension and diabetes, but their evidence came from a prospective cohort study in two Kenyan counties rather than a national household sample and focused on financial risk protection rather than ongoing treatment use [7]. Our cross-sectional national estimate complements that work by showing that the weak adjusted insurance association with current medication use is not unique to selected counties. A recent household fixed-effects study from India likewise found no significant improvement across hypertension-care cascade steps with insurance coverage alone [30]. It also differs in focus from the 2015 Kenya STEPs analysis by Oyando et al., which examined pro-rich inequity in hypertension screening and treatment and used decomposition methods rather than a diagnosed-adult insurance and treatment-cascade framework [31]. The findings also complement the earlier Kenya DHS literature by Kimani et al. and Kazungu and Barasa, which documented low and unequal insurance coverage in the general population [4, 5].

The inequality results are especially revealing. Insurance coverage was strongly pro-rich, with a pooled concentration index for any insurance broadly consistent with wealth-skewed patterns previously documented in Kenya and across sub-Saharan Africa [5, 6]. Yet the concentration index for treatment gaps was close to zero and unchanged in direction under the Erreygers-corrected sensitivity analysis [15, 16]. Interpreted against concentration-index methodology for bounded outcomes, that contrast suggests that untreated chronic disease was not confined to the poorest households alone. Wealth still shaped the probability of being insured, but once respondents had a diagnosed condition, gaps in ongoing treatment extended across the socioeconomic distribution. In other words, while wealth-related inequity remained concentrated in insurance enrolment, the burden of untreated diagnosed disease was distributed broadly across wealth groups, consistent with the inequality estimates reported above.

Sex differences add another layer. Men had higher insurance coverage than women, but women had larger treatment gaps and reported substantially greater financial (money-for-treatment) and distance barriers when uninsured. The DHS access-barrier items used here map closely to the three-delays framework described by Thaddeus and Maine [32] and to broader evidence on barriers to healthcare access among women in sub-Saharan Africa [33]. Our findings add a chronic-disease dimension to that literature: the uninsured faced a heavier burden of practical barriers even within a diagnosed NCD population. This is relevant for SHA implementation because enrolment drives that do not account for gendered barriers to cash access and service navigation may leave a substantial treatment deficit intact.

The SHA transition makes these findings timely rather than historical. KDHS 2022 is the last nationally representative survey before the 1 October 2024 financing reform, so it provides a baseline against which SHA should be judged [3]. If SHA succeeds only in expanding enrolment while leaving medicine availability, benefit-package design, reimbursement reliability, and frontline chronic-care delivery unchanged, then treatment gaps of the magnitude documented here may persist. Facility, household, and benefits-package evidence alike show persistent weaknesses in NCD readiness, medicine access, and chronic-care priority setting [34–38]. Patient-cost evidence points in the same direction: treatment can remain financially burdensome even within public-sector care [39]. Evidence from Kenya’s purchasing and UHC reform literature suggests that enrolment expansion must be matched by credible financing and benefit-design reforms [22, 23, 40, 41]. As Kenya institutionalises SHA-era benefits-package priority setting, outpatient NCD medicines and reliable reimbursement for chronic care should remain central implementation tests [37].

This study has limitations. The design was cross-sectional, so associations should not be interpreted causally and temporal ordering between current insurance and current medication use cannot be established. Diagnoses were captured through respondent-reported prior clinician diagnosis rather than clinical reconfirmation; while this is consistent with practice in DHS- and STEPS-based cascade studies of hypertension and diabetes in low- and middle-income countries [12, 13], such measures can underestimate true prevalence and may be patterned by socioeconomic status (e.g., access to clinical contact), as documented in the WHO Study on global AGEing and adult health [43]. Medication use and roster-based insurance coverage were similarly respondent-reported, which may introduce recall or reporting error and some merge-related misclassification. The women’s analysis was restricted to the long-questionnaire subsample and to ages 18-49 years, while men were observed to age 54 years, so the sex-stratified samples are not perfectly equivalent. Because hypertension, diabetes, and care-seeking all vary with age, excluding women aged 50-54 years likely makes the female analytic sample younger than the corresponding male sample and may shift women’s coverage and treatment estimates relative to the full adult female diagnosed population; the direction for treatment gaps is uncertain, so sex contrasts should be interpreted descriptively rather than as direct population-equivalent comparisons. Residual confounding is also likely because insurance enrolment may be correlated with unmeasured health-seeking behaviour, disease severity, or provider contact. Finally, the treatment gap was defined as not currently taking medication for at least one diagnosed condition, meaning partially treated individuals with multiple diagnoses are classified as having a gap. While clinically appropriate for complete chronic-care coverage, this produces higher estimates than a stricter no-medication-for-any-diagnosed-condition definition. Reassuringly, the stricter sensitivity analysis yielded similar prevalence estimates and insurance APRs (Additional file 3: Supplementary Table S2). Because male interviews were conducted in a household subsample, pooled estimates required a male-subsample correction and should be interpreted as secondary to the sex-stratified analyses.

Within the pooled comorbid subgroup, 25 respondents had a gap in one condition and 24 had gaps in both, underscoring why the inclusive treatment-gap definition matters for interpretation.

## Conclusions

Among Kenyan adults who reported diagnosed hypertension or diabetes in KDHS 2022, insurance coverage remained incomplete and treatment gaps remained large at the end of the NHIF era. Insurance coverage was strongly pro-rich, but treatment gaps were widespread across wealth groups, indicating a chronic-care problem that extended beyond enrolment inequity alone. Women had larger treatment gaps than men and, when uninsured, were substantially more likely to report money for treatment and distance as serious barriers to care. As Kenya implements SHA, these findings suggest that universal enrolment targets should be paired with deeper financial protection, reliable medicine access, and benefit packages designed to reach adults already living with diagnosed conditions.

## Supporting information

Odds-ratio sensitivity analyses are reported in Supplementary Table S1

We repeated the prevalence and regression analyses using the stricter no-treatment definition and report those results in a separate file (Additional

and the stricter sensitivity analysis (a respondent classified as having a gap only when no medication was taken for any diagnosed condition) is repor

## Data Availability

The 2022 Kenya Demographic and Health Survey microdata analysed in this study are not openly available; they were obtained from The DHS Program following registration and approval of a standard data-access application, and may be obtained by other researchers through the same procedure at https://dhsprogram.com. All analysis scripts and derived non-identifiable summary outputs are publicly available without restriction at https://github.com/gondamol/Kenya_DHS_Studies.

https://dhsprogram.com/data/dataset/Kenya_Standard-DHS_2022.cfm

https://github.com/gondamol/Kenya_DHS_Studies

## Abbreviations

APR: adjusted prevalence ratio
CI: confidence interval
DM: diabetes mellitus
DHS: Demographic and Health Survey
HTN: hypertension
KDHS: Kenya Demographic and Health Survey
NHIF: National Health Insurance Fund
NCD: non-communicable disease
OR: odds ratio
SHA: Social Health Authority

## Declarations

### Ethics approval and consent to participate

This study used de-identified secondary data from the 2022 Kenya Demographic and Health Survey, obtained from The DHS Program under authorisation letter reference 220623 for the registered project titled “Impact of NHIF Enrollment on Financial Catastrophic Health Expenditure”, authorised on 20 October 2025. The original survey received ethical approval from the Kenya Medical Research Institute Scientific and Ethics Review Unit (KEMRI SERU; KEMRI/SERU) and the ICF Institutional Review Board (ICF IRB reference: 222125.0.000 / FWA00000845; The DHS Program standard approval covering KDHS 2022, as confirmed in the KDHS 2022 final report). No new participant contact occurred in this secondary analysis.

### Consent for publication

Not applicable.

### Availability of data and materials

The datasets analysed during the current study are available from The DHS Program (www.dhsprogram.com) following registration and approval of the standard data access application. Study-specific analysis scripts are available at https://github.com/gondamol/Kenya_DHS_Studies.

### Authors’ contributions

NWA conceptualised the study, conducted data analysis, and drafted the manuscript. JO provided critical review, methodological inputs, and substantive revisions that strengthened the framing, analysis, and interpretation of the manuscript. HFH contributed to study design, interpretation of findings, and critical revision of the manuscript. All authors reviewed and approved the final version.

## Acknowledgements

The authors thank The DHS Program for access to KDHS 2022 microdata.

