## Supplementary material for "Health insurance coverage, affordability barriers, and treatment gaps among Kenyan adults with diagnosed hypertension or diabetes before the Social Health Authority transition: a sex-stratified analysis of the 2022 Kenya Demographic and Health Survey": Odds-ratio sensitivity analyses are reported in Supplementary Table S1

### STROBE checklist

Study type: Cross-sectional secondary analysis of KDHS 2022.

| Item | Reporting element | Location in manuscript |
| --- | --- | --- |
| 1 | Study design identified in title and abstract | Title; Abstract |
| 2 | Background and rationale | Background |
| 3 | Objectives and prespecified questions | End of Background |
| 4 | Key elements of study design | Methods: Study design and data source |
| 5 | Setting, survey context, and dates | Methods: Study design and data source |
| 6 | Eligibility criteria and participant selection | Methods: Study population |
| 7 | Definitions of outcomes, exposures, predictors, and effect modifiers | Methods: Outcome definition; Insurance exposure and covariates |
| 8 | Data sources and measurement | Methods: Analytic files and merge strategy; Outcome definition; Insurance exposure and covariates |
| 9 | Potential sources of bias | Discussion: Limitations |
| 10 | Study size | Methods: Study population; Results |
| 11 | Handling of quantitative variables | Methods: Insurance exposure and covariates; Survey weighting and statistical analysis |
| 12 | Statistical methods, confounding control, subgroup analyses, interaction tests, and sensitivity analyses | Methods: Survey weighting and statistical analysis |
| 13 | Participant flow and analytic sample | Methods: Study population; Results; Table 1 |
| 14 | Descriptive characteristics of participants | Results: Sample characteristics and insurance profile; Table 1 |
| 15 | Outcome data | Results: Treatment cascade by diagnosis profile; Wealth-related inequality and women-specific access barriers; Tables 2-5; Figures 1-3 |
| 16 | Main results with adjusted and unadjusted estimates | Results: Adjusted associations with treatment gaps; Tables 2-5 |
| 17 | Other analyses, including interaction and sensitivity analyses | Results: Adjusted associations with treatment gaps; Table 5; Supplementary Table S1 |
| 18 | Key findings in relation to study objectives | Discussion: opening paragraph |
| 19 | Study limitations | Discussion: Limitations |
| 20 | Interpretation considering objectives, limitations, and comparable evidence | Discussion |
| 21 | Generalisability | Discussion: Limitations; Conclusions |
| 22 | Funding and role of funders | Front matter; Declarations: Funding |
