## Supplementary material for "Health insurance coverage, affordability barriers, and treatment gaps among Kenyan adults with diagnosed hypertension or diabetes before the Social Health Authority transition: a sex-stratified analysis of the 2022 Kenya Demographic and Health Survey": We repeated the prevalence and regression analyses using the stricter no-treatment definition and report those results in a separate file (Additional

**Supplementary Table S1. Odds-ratio sensitivity analyses for the treatment gap among diagnosed adults, Kenya DHS 2022.**

| **Covariate** | **Women OR (95% CI)** | **Men OR (95% CI)** | **Pooled OR (95% CI)** |
| --- | --- | --- | --- |
| Insurance |  |  |  |
| Uninsured (ref=1.00) | 1.00 (ref) | 1.00 (ref) | 1.00 (ref) |
| Insured | 0.75 (0.52, 1.08) | 0.80 (0.43, 1.50) | 0.78 (0.56, 1.10) |
| Age (per year) | 0.95 (0.93, 0.98) | 0.96 (0.93, 0.99) | 0.96 (0.94, 0.98) |
| Diagnosis |  |  |  |
| Both HTN+DM (ref=1.00) | 1.00 (ref) | 1.00 (ref) | 1.00 (ref) |
| HTN only | 1.69 (0.49, 5.80) | 4.18 (1.83, 9.51) | 2.92 (1.49, 5.72) |
| DM only | 0.62 (0.16, 2.40) | 0.61 (0.20, 1.91) | 0.65 (0.28, 1.51) |
| Wealth |  |  |  |
| Richest (ref=1.00) | 1.00 (ref) | 1.00 (ref) | 1.00 (ref) |
| Richer | 0.77 (0.47, 1.24) | 0.87 (0.38, 1.98) | 0.82 (0.52, 1.31) |
| Middle | 0.82 (0.48, 1.41) | 0.66 (0.26, 1.66) | 0.74 (0.45, 1.22) |
| Poorer | 0.82 (0.44, 1.54) | 0.84 (0.29, 2.42) | 0.86 (0.49, 1.52) |
| Poorest | 0.62 (0.29, 1.33) | 0.94 (0.29, 3.00) | 0.79 (0.40, 1.54) |
| Education |  |  |  |
| None (ref=1.00) | 1.00 (ref) | 1.00 (ref) | 1.00 (ref) |
| Primary | 0.58 (0.27, 1.26) | 0.52 (0.12, 2.23) | 0.50 (0.23, 1.08) |
| Secondary+ | 0.69 (0.30, 1.56) | 0.49 (0.12, 2.03) | 0.54 (0.24, 1.20) |
| Residence |  |  |  |
| Rural (ref=1.00) | 1.00 (ref) | 1.00 (ref) | 1.00 (ref) |
| Urban | 0.92 (0.59, 1.43) | 0.92 (0.47, 1.83) | 0.94 (0.64, 1.38) |
| Employment |  |  |  |
| Agriculture (ref=1.00) | 1.00 (ref) | 1.00 (ref) | 1.00 (ref) |
| Professional/clerical | 0.74 (0.43, 1.25) | 0.67 (0.28, 1.59) | 0.74 (0.46, 1.21) |
| Sales/services | 0.52 (0.29, 0.93) | 0.39 (0.17, 0.87) | 0.47 (0.30, 0.73) |
| Manual | 1.19 (0.62, 2.29) | 0.52 (0.26, 1.05) | 0.77 (0.48, 1.24) |
| Not working | 0.66 (0.41, 1.06) | 0.32 (0.12, 0.87) | 0.54 (0.34, 0.85) |
| Province |  |  |  |
| Central (ref=1.00) | 1.00 (ref) | 1.00 (ref) | 1.00 (ref) |
| Coast | 0.53 (0.28, 0.99) | 0.36 (0.15, 0.88) | 0.47 (0.28, 0.80) |
| Eastern | 0.76 (0.44, 1.32) | 0.78 (0.31, 1.96) | 0.81 (0.50, 1.31) |
| Nairobi | 0.41 (0.19, 0.89) | 0.69 (0.18, 2.69) | 0.53 (0.25, 1.10) |
| NE | 0.64 (0.28, 1.46) | 0.28 (0.07, 1.17) | 0.38 (0.17, 0.84) |
| Nyanza | 0.44 (0.25, 0.75) | 1.23 (0.55, 2.76) | 0.74 (0.46, 1.18) |
| Rift Valley | 0.72 (0.44, 1.19) | 0.65 (0.29, 1.47) | 0.74 (0.48, 1.15) |
| Western | 0.61 (0.34, 1.10) | 0.39 (0.15, 1.00) | 0.52 (0.30, 0.90) |

Source: Kenya DHS 2022. Survey-weighted estimates. Directional conclusions were unchanged from the adjusted prevalence-ratio models. OR = odds ratio; CI = confidence interval.
