## Supplementary material for "Health insurance coverage, affordability barriers, and treatment gaps among Kenyan adults with diagnosed hypertension or diabetes before the Social Health Authority transition: a sex-stratified analysis of the 2022 Kenya Demographic and Health Survey": and the stricter sensitivity analysis (a respondent classified as having a gap only when no medication was taken for any diagnosed condition) is repor

**Supplementary Table S2. Sensitivity analysis using a stricter no-medication-for-any-diagnosed-condition definition, Kenya DHS 2022.**

| **Outcome definition** | **Women prevalence % (95% CI)** | **Men prevalence % (95% CI)** | **Pooled prevalence % (95% CI)** | **Women APR for insured vs uninsured (95% CI)** | **Men APR for insured vs uninsured (95% CI)** | **Pooled APR for insured vs uninsured (95% CI)** |
| --- | --- | --- | --- | --- | --- | --- |
| Main analysis: not on medication for at least one diagnosed condition | 67.1 (63.5, 70.6) | 59.8 (53.8, 65.8) | 63.8 (60.4, 67.3) | 0.91 (0.81, 1.03) | 0.93 (0.75, 1.16) | 0.93 (0.83, 1.04) |
| Sensitivity analysis: no medication for any diagnosed condition | 65.8 (62.0, 69.7) | 57.8 (51.7, 63.8) | 62.2 (58.7, 65.7) | 0.93 (0.83, 1.04) | 0.93 (0.75, 1.15) | 0.94 (0.85, 1.04) |

Note: The main analysis defined the treatment gap as not currently taking medication for at least one diagnosed condition. The sensitivity analysis classified respondents as having a gap only when they reported no medication for any diagnosed condition. APR = adjusted prevalence ratio; CI = confidence interval.
